# The Impact of Post Embryo Transfer SARS-CoV-2 Infection on Pregnancy in In Vitro Fertilization: A Prospective Cohort Study

**DOI:** 10.1101/2023.07.25.23293116

**Authors:** Xue-Fei Li, Yong-Jia Zhang, Ying-Ling Yao, Ming-Xing Chen, Li-Li Wang, Meng-Di Wang, Xin-Yue Hu, Xiao-Jun Tang, Zhao-Hui Zhong, Li-Juan Fu, Xin Luo, Xing-Yu Lv, Li-Hong Geng, Qi Wan, Yu-Bin Ding

## Abstract

**Importance:** Limited knowledge exists on the effects of SARS-CoV-2 infection after embryo transfer, despite an increasing number of studies exploring the impact of previous SARS-CoV-2 infection on IVF outcomes.

**Objective:** This prospective cohort study aimed to assess the influence of SARS-CoV-2 infection at various time stages after embryo transfer on pregnancy outcomes in patients undergoing conventional in vitro fertilization/intracytoplasmic sperm injection-embryo transfer (IVF/ICSI) treatment.

**Design:** The study was conducted at a single public IVF center in China.

**Setting:** This was a population-based prospective cohort study.

**Participants:** Female patients aged 20 to 39 years, with a body mass index (BMI) between 18 and 30 kg/m^2^, undergoing IVF/ICSI treatment, were enrolled from September 2022 to December 2022, with follow-up until March 2023.

**Exposure:** The pregnancy outcome of patients was compared between those SARS-CoV-2-infected after embryo transfer and those noninfected during the follow-up period.

**Main Outcomes and Measures:** The pregnancy outcomes included biochemical pregnancy rate, implantation rate, clinical pregnancy rate, and early miscarriage rate.

**Results:** A total of 857 female patients undergoing IVF/ICSI treatment were included in the analysis. We observed the incidence of SARS-CoV-2 infection within 10 weeks after embryo transfer. The biochemical pregnancy rate and implantation rate were lower in the infected group than the uninfected group (58.1% vs 65.9%; 36.6% vs 44.0%, respectively), but no statistically significant. Although, the clinical pregnancy rate was significant lower in the infection group when compared with the uninfected group (49.1%vs 58.2%, p < 0.05), after adjustment for confounders, this increased risk was no longer significant between the two groups (adjusted OR, 0.736, 95% CI, 0.518-1.046). With continued follow-up, a slightly higher risk of early miscarriage in the infected group compared to the uninfected group (9.3% vs 8.8%), but it was not significant (adjusted OR, 0.907, 95% CI, 0.414-1.986).

**Conclusions and Relevance:** The study’s findings suggested that SARS-CoV-2 infection within 10 weeks after embryo transfer may have not significantly affect pregnancy outcomes. This evidence allays concerns and provides valuable insights for assisted reproduction practices.

**Key points:** *Question:* Did the infection of severe acute respiratory syndrome coronavirus 2 (SARS-CoV-2) after embryo transfer affect pregnancy outcomes?

*Findings:* In this prospective cohort study involving 857 patients, we made a pioneering discovery that SARS-CoV-2 infection following embryo transfer did not exhibit adverse impact on the biochemical pregnancy rate, embryo implantation rate, clinical pregnancy rate, and early miscarriage rate.

*Meaning:* The evidence from this study alleviates existing concerns and offers new insights into the actual risk of SARS-CoV-2 infection after embryo transfer in assisted reproduction.

## Introduction

The COVID-19 pandemic caused by severe acute respiratory syndrome coronavirus 2 (SARS-CoV-2) has had a profound impact on human health and life, affecting billions of individuals worldwide^1,2^. Additionally, concerns have arisen regarding the potential effects of SARS-CoV-2 on human fertility, gametes, pregnancy, and assisted reproduction techniques^3,4^. Numerous studies have demonstrated the susceptibility of both male and female reproductive systems to SARS-CoV-2 infection, as they possess the essential proteins and receptors for viral cell entry^5-8^. However, the impact of SARS-CoV-2 infection on semen parameters and male reproductive potential remains a topic of controversy. Women who have experienced severe SARS-CoV-2 infection have documented transient changes in their menstrual patterns^8,9^. However, no significant impact of COVID-19 on ovarian reserves, ovarian function or follicular fluid parameters has been reported^10,11^. It should be noted that while the endometrium is highly susceptible to SARS-CoV-2 infection, it remains unclear whether this infection can alter uterine receptivity and embryo implantation.

During the COVID-19 pandemic, lifestyle changes and infections can potentially affect the success of assisted reproductive technology (ART) treatment for couples^12^. For females infected with SARS-CoV-2 before oocyte retrieval in ART treatment, there is no evidence suggesting adverse effects on cycle and pregnancy outcomes^9,13-16^. However, previous SARS-CoV-2 infection with oocyte retrieval before infection showed a slight decrease in blastocyst formation rate and long-term negative effects on oocyte yield. Patients with retrieval more than 180 days after SARS-CoV-2 infection exhibited decreased pregnancy rates, especially when recovered less than 60 days before embryo transfer in ART cycles. ^17-19^. Currently, there are no investigations into whether post-embryo transfer SARS-CoV-2 infection adversely influences pregnancy outcomes in IVF data.

In December 2022, China faced a momentous shock when nearly a billion people were affected by severe acute SARS-CoV-2 infection, primarily driven by the BA.5 and BF.7 Omicron subvariants. This outbreak came in the wake of the unexpected termination of the “Zero-COVID” policy, which had successfully kept Chinese citizens largely free from adverse infections for the preceding three years. Amidst this situation, fertility and in vitro fertilization (IVF) centers observed a rise in the number of infected patients, creating an opportunity to investigate the impact of post-embryo transfer SARS-CoV-2 infection on pregnancy outcomes in female patients who received IVF/ICSI-ET therapy^14,20^. This study was based on data collected from women undergoing ART treatment in China between September 2022 and December 2022, as part of the CYART prospective cohort. The new findings hold significant implications for healthcare professionals and patients seeking ART treatment amidst the ongoing COVID-19 pandemic. Furthermore, our research contributes to the establishment of evidence-based guidelines for IVF treatment in these challenging circumstances, as it provides a deeper understanding of how post-embryo transfer SARS-CoV-2 infection may influence pregnancy outcomes.

## Methods

### Study population and design

The CYART cohort was established on September 1, 2021, at Sichuan Jinxin Xinan Women & Children’s Hospital in southwest China, with the primary objective of investigating factors related to infertility and ART treatments. The study received approval from the Ethics Committee of Sichuan Jinxin Xinan Women & Children’s Hospital (No. 2021014) and the Ethics Committee of Chongqing Medical University (No. 2021060). Patient characteristics and information regarding IVF treatment were extracted from the center’s electronic medical records. A total of 3,867 people received ART treatment at the center, the exclusion criteria as follows: (1) age <20 or >40 years; (2) BMI <18 or >30 kg/m^2^; (3) previous SARS-CoV-2 infection before embryo transfer; (4) receipt of donated oocytes; (5) absence of oocyte retrieval, complete embryo cryopreservation, or canceled embryo transfer; (6) lost to follow-up. Finally, 857 patients met the research criteria. They received ART therapy from September 2022 to December 2022, with follow-up until March 2023. This period coincided with the largest surge of COVID-19 cases in China following the termination of the “Zero-COVID” policy. During the study, a questionnaire regarding SARS-CoV-2 infection was administered to the patients through on-site or telephone surveys, and relevant follow-up was conducted (Supplementary Appendix). The diagnosis of SARS-CoV-2 infection in patients was based on nucleic acid testing or antigen self-testing, with the addition of patient-reported symptoms. All patients included in the study provided written informed consent.

In order to assess the potential impact of the time interval between embryo transfer and reporting of SARS-CoV-2 infection on pregnancy outcomes, we categorized the patients into three stages: (1) ≤14 days, (2) ≤28 days, and (3) ≤10 weeks (Our study focused on SARS-CoV-2 infection after embryo transfer. Since early miscarriage usually occurs within the first 12 weeks of pregnancy, the first two weeks may not be an infection that occurs after embryo transfer. Therefore, we refer to this period collectively as “10 weeks after embryo transfer” rather than “12 weeks of gestation.”). These stages correspond to key pregnancy outcomes, including chemical pregnancy, clinical pregnancy, and early miscarriage (≤12 weeks of pregnancy). The flow of study participants was depicted in Figure 1. Day1 was defined as the day after embryo transfer. Patients reporting SARS-CoV-2 infection between Day1 and Day14 were assigned to group A (n=136), while those remaining uninfected were assigned to group B (n=721). The primary outcome in this first stage was biochemical pregnancy rate. For uninfected patients from the first stage, we continued to follow up until Day 28. During this period, patients reporting SARS-CoV-2 infection were divided into subgroup B1 (n=31), while the remaining uninfected patients were assigned to subgroup B2 (n=686). In this second stage, the infection group consisted of group A combined with subgroup B1(n=171), and the uninfected group consisted of subgroup B2 (n=686). The primary outcome was clinical pregnancy rate, and the secondary was implantation rate. Follow-up continued until 10 weeks after embryo transfer. Patients reporting SARS-CoV-2 infection during this period were assigned to subgroup C1 (n=525), while uninfected patients were assigned to subgroup C2 (n=161). This represented the third stage, the infection group consisted of group A combined with subgroup B1 and subgroup C1 (n=691), and the uninfected group consisted of subgroup C2 (n=161). The observed outcome measure was early miscarriage rate.

**Figure 1.**
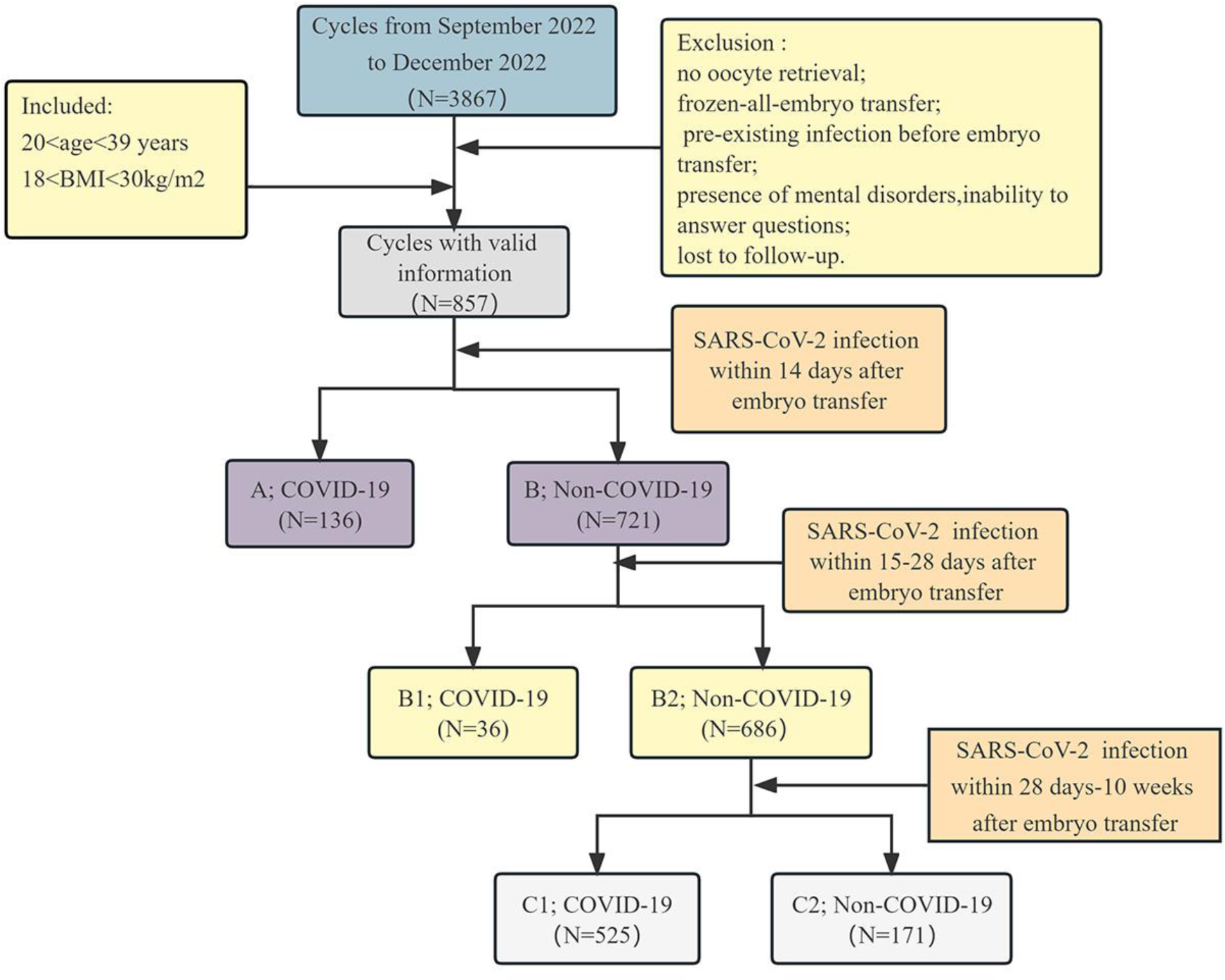
flowchart of the study.

### IVF/ICSI-ET protocols

The controlled ovarian stimulation (COS) protocols in our center mainly includes gonadotropin-releasing hormone (GnRH) antagonist protocol, long follicular phase protocol, and other protocols, including mild stimulation and luteal phase stimulation protocols. Details of the COS protocols have been explained in detail in previous studies^21,22^. When ultrasound indicates that 2-3 follicles greater than 18 mm were given human chorionic gonadotrophin (HCG) or gonadotropin releasing hormone agonist (GnRH-a) combined trigger. Egg-retrieval surgery was performed 36-38 hours later, followed by fresh embryo transfer on the third or fifth day after the surgery.

### Outcome measures and definitions

We focused on evaluating the impact of SARS-CoV-2 infection on pregnancy outcomes in patients undergoing IVF/ICSI treatment after embryo transfer. The primary outcomes included biochemical pregnancy rate, clinical pregnancy rate, and early miscarriage rate, and secondary outcome was implantation rate. Biochemical pregnancy was defined as the serum β-HCG > 25U/L 14 days after embryo transfer. Clinical pregnancy was confirmed through transvaginal ultrasound 28 days after embryo transfer, with the presence of a gestational sac or fetal heart being indicative of a clinical pregnancy. Implantation rate was calculated as the number of intrauterine gestational sacs divided by the number of embryos transferred. Early miscarriage was defined as the number of spontaneous miscarriages occurring within 12 weeks of gestation to the number of clinical pregnancies.

### Statistical Analysis

We performed all analyses using SPSS software (version 26.0, IBM, USA). Since none of the continuous variables was normally distributed using Shapiro-Wilk normality test, the continuous variables were presented as medians (IQR); while Categorical variables were expressed as number (n) and percentage (%). Mann-Whitney U test or Student’s t-tests were used for continuous variables and the Chi-square test was used for categorical variables. Multivariate logistic regression was used to calculate each outcome’s adjusted odds ratios and 95% CI. All statistical tests were two-sided, and P < 0.05 was considered to be statistically significant.

## Results

### Baseline Characteristics

In the first stage, patients contracting SARS-CoV-2 infection within 14 days after embryo transfer were divided into two groups: infected (136 patients) and uninfected (721 patients) (Table 1). Their had similar results in terms of female age, BMI, infertility durations, hormone levels, infertility cause, type, COS protocol, and transferred embryos. However, significant differences were observed in fertilization type, transferred embryo type, and number of high-quality embryos. The uninfected group had a higher percentage of IVF treatment (87.5% vs 81.0%) and a greater proportion underwent split-stage embryo transfer (73.5% vs. 62.7%) with more embryos transferred.

**Table 1.**
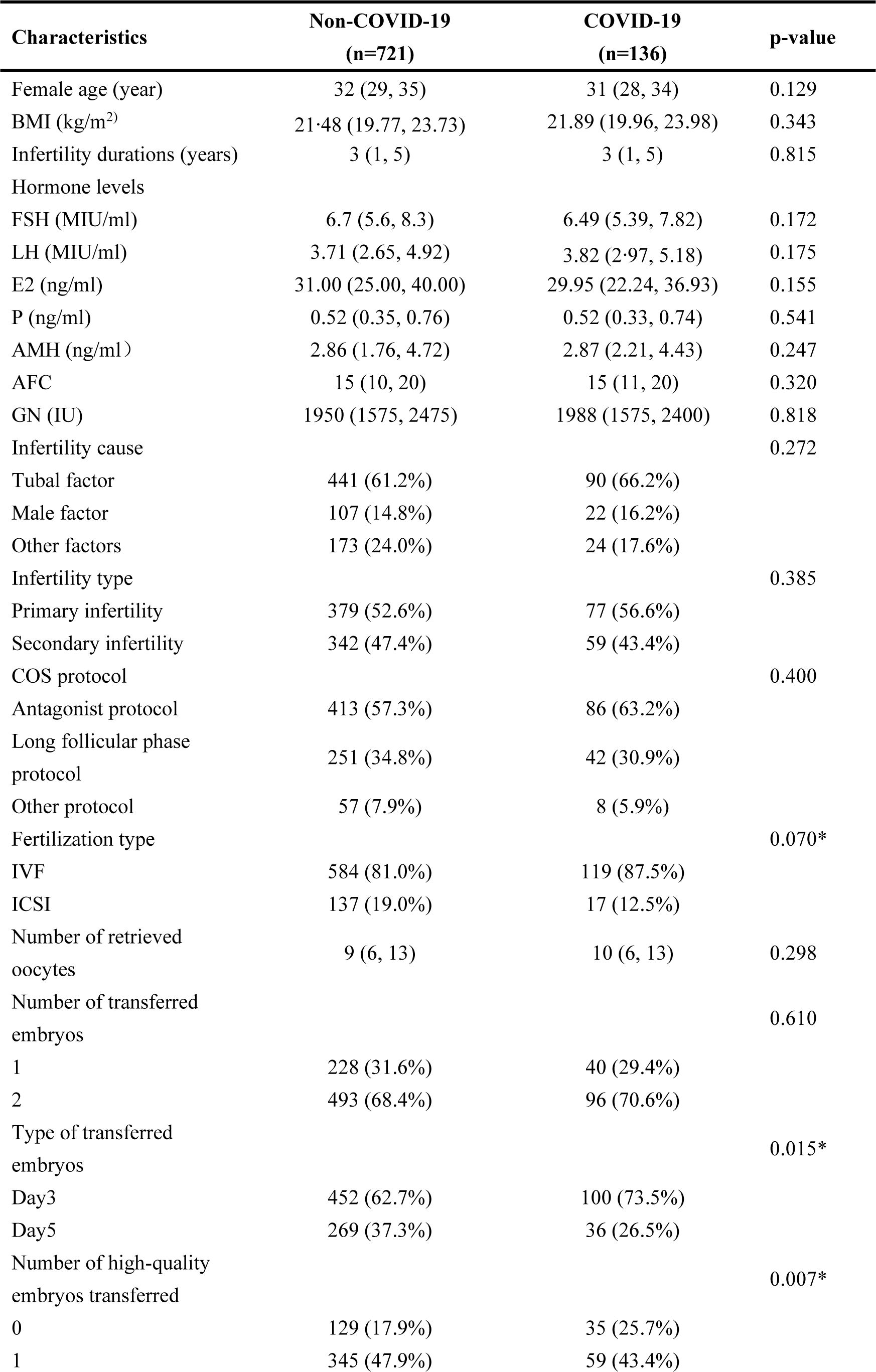
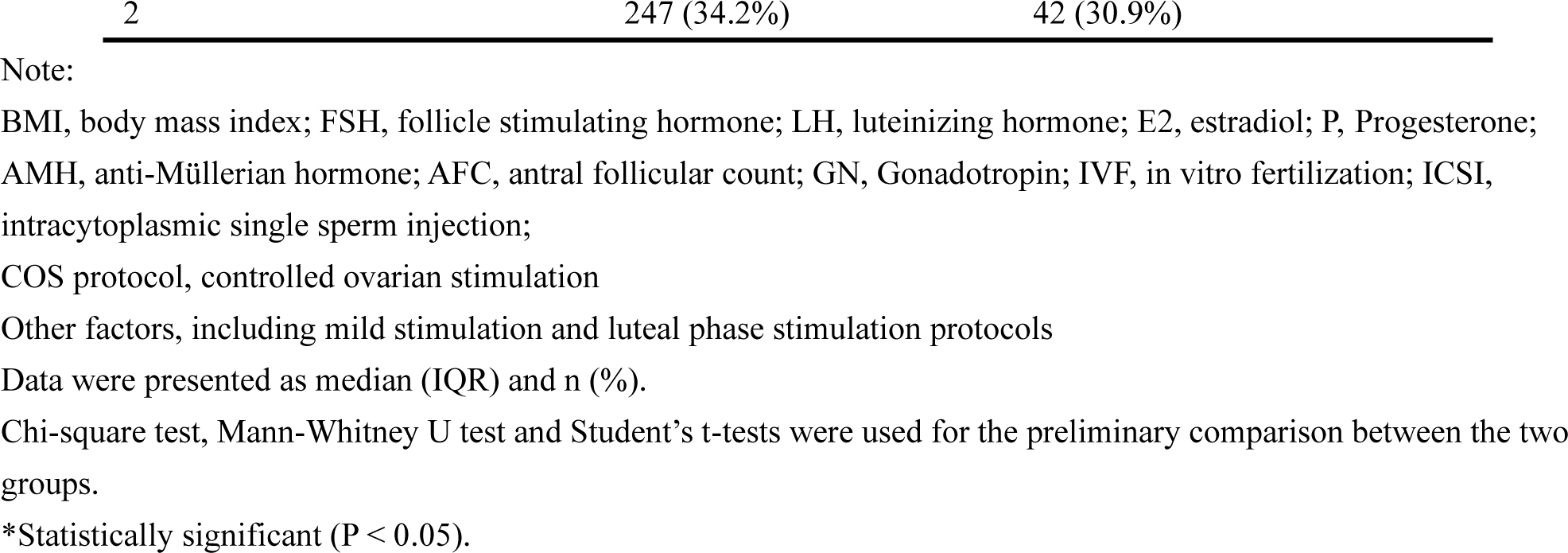
Baseline characteristics of the infected and uninfected patients within 14 days of embryo transfer.

In the second stage, patients were divided based on infection within 28 days after embryo transfer, with 171 in the infected group and 686 in the uninfected group (Table 2). Baseline characteristics showed no statistically significant differences in female age, BMI, hormone levels, infertility durations, infertility cause, infertility type, COS protocol, and number of transferred embryos. However, significant differences were observed in fertilization type, type of transferred embryos, and number of high-quality embryos transferred. The infected group had a higher percentage of patients undergoing IVF treatment compared to the uninfected group (88.3% vs. 80.5%), and a higher proportion of patients in the infected group underwent transplantation during the cleavage stage (74.9% vs. 61.7%).

**Table 2.**
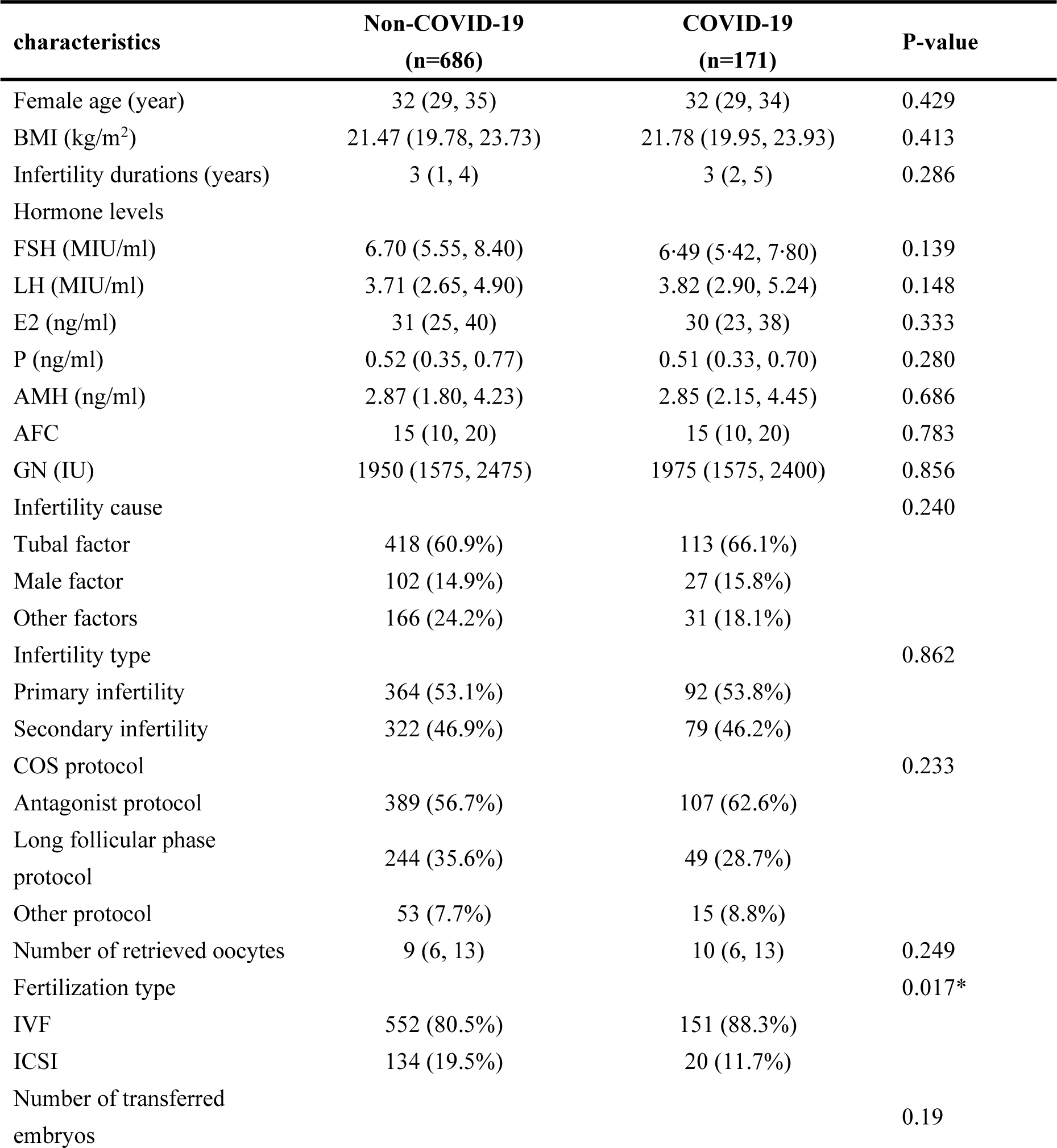
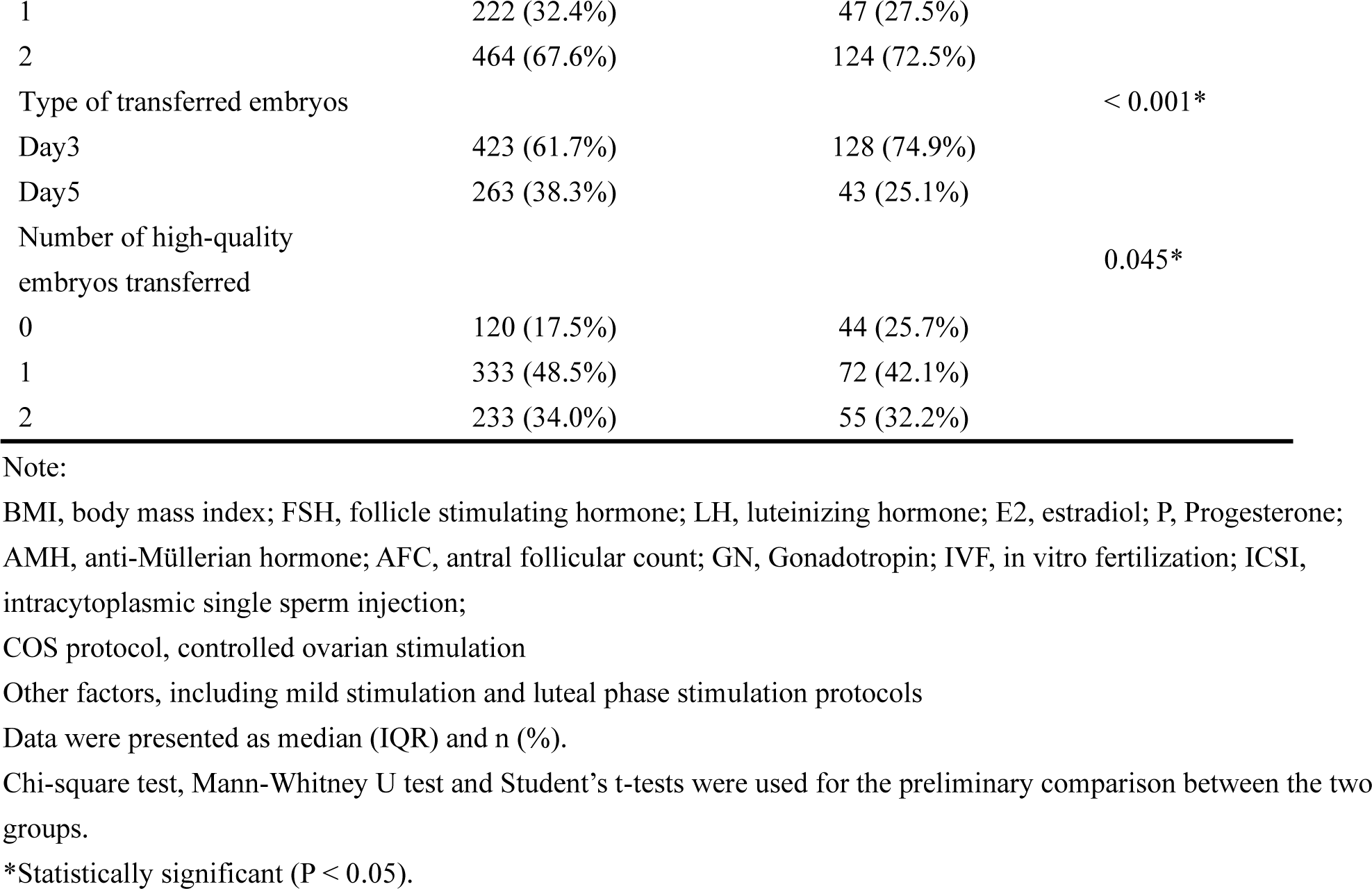
Baseline characteristics of infected and uninfected patients within 28 days of embryo transfer.

In the third stage, patients were categorized into two groups based on SARS-CoV-2 infection within 10 weeks after embryo transfer. The infection group comprised 691 patients, and the uninfected group consisted of 161 patients (Table 3). There were no significant differences between the two groups in terms of female age, BMI, LH, E2, P, AFC, GN, infertility durations, infertility cause, infertility type, fertilization type, number of transferred embryos, type of transferred embryo, and number of high-quality embryos transferred. However, statistically significant differences were observed in FSH, AMH, and COS protocol. The uninfected group had higher levels of FSH (7.04 vs. 6.57) and AMH (3.19 vs. 2.84). Additionally, a lower proportion of patients in the uninfected group used the antagonist protocol for ovulation induction compared to the infection group (36.6% vs. 62.9%). Furthermore, a higher number of patients in the uninfected group used the long follicular phase protocol for ovulation induction (57.2% vs. 29.2%).

**Table 3.**
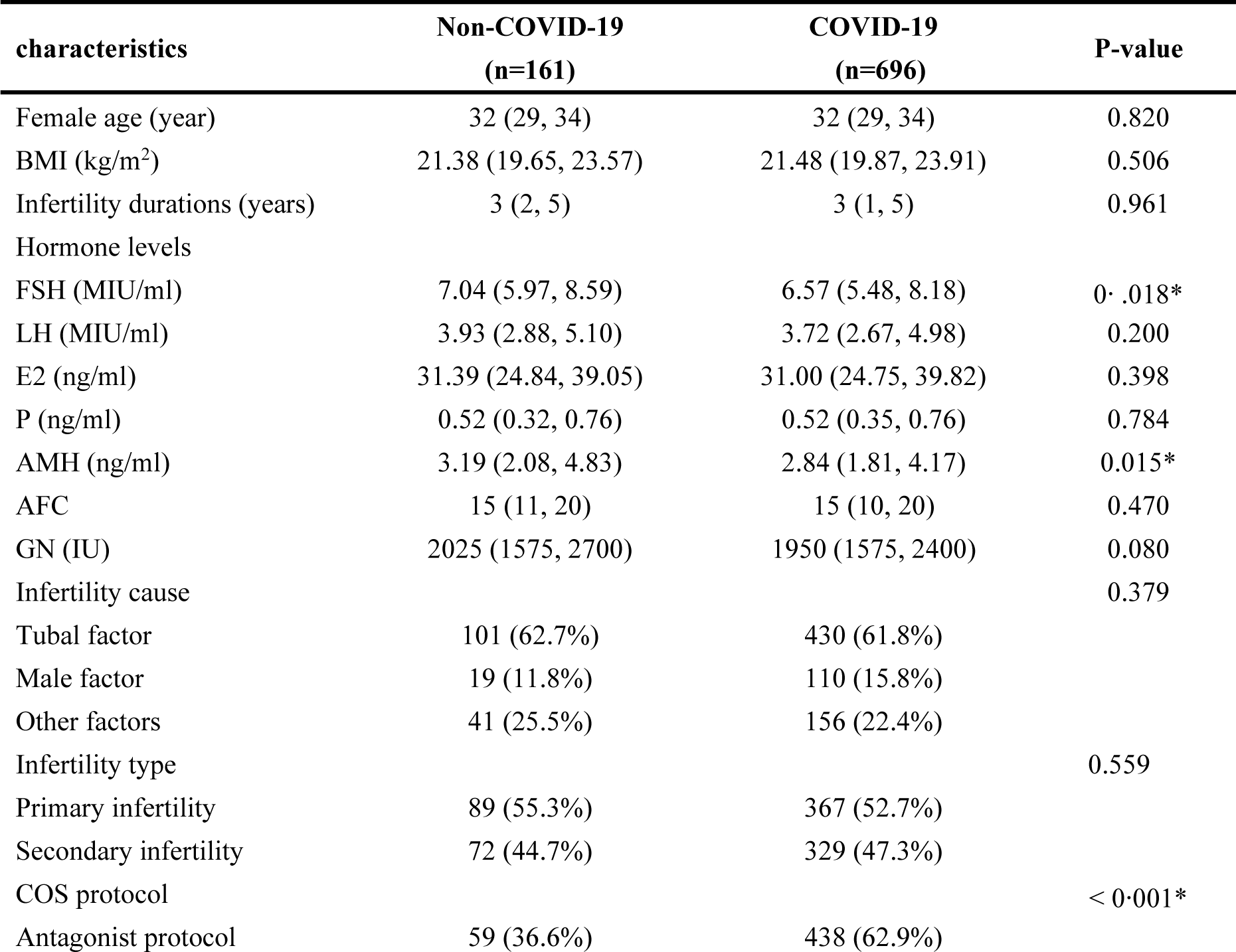
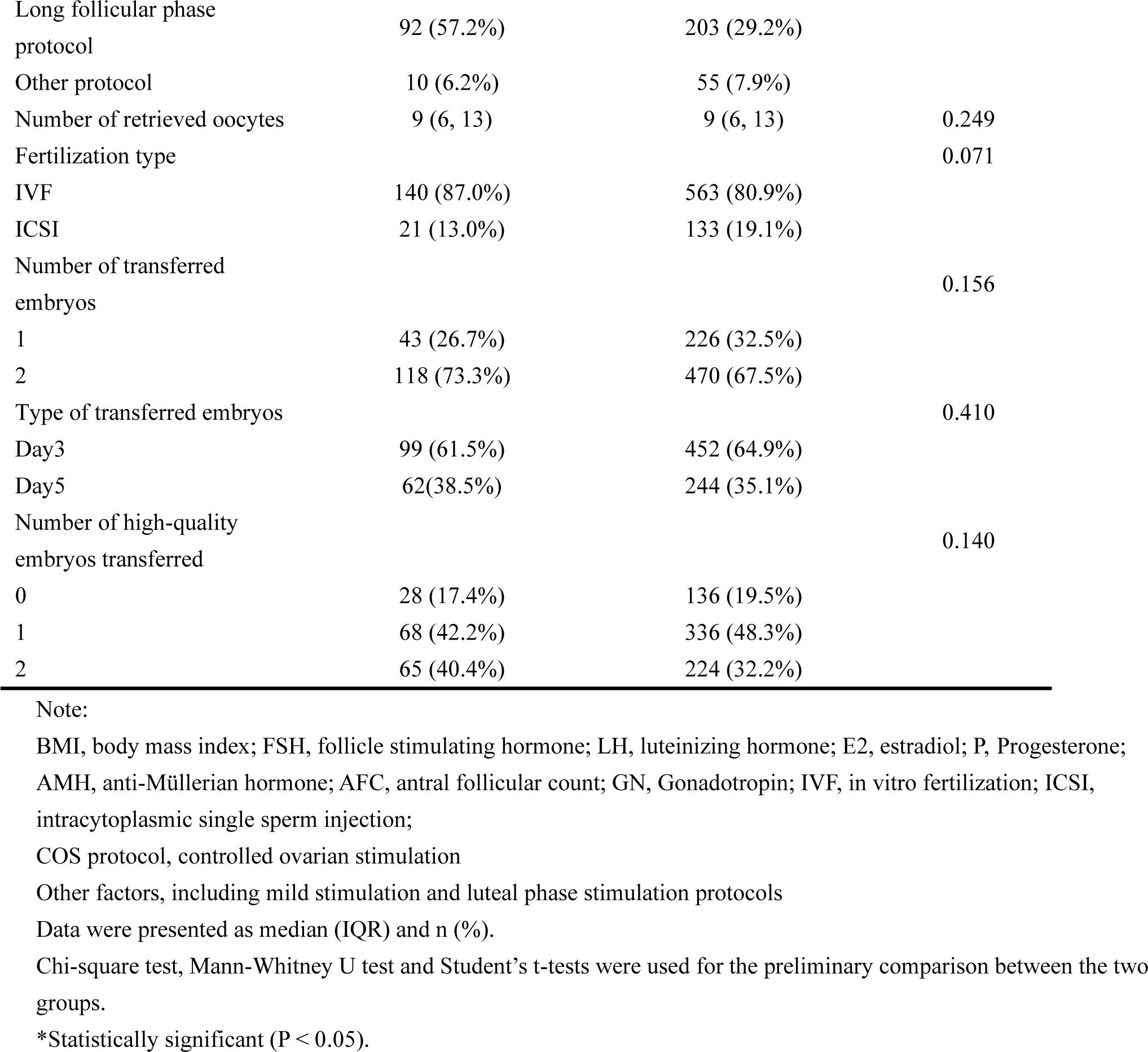
Baseline characteristics of infected and uninfected patients within 10 weeks of embryo transfer.

### Comparison of pregnancy outcome

Pregnancy outcomes were presented in Table 4, and multivariate logistic regression was conducted to minimize the imbalance of baseline characteristics (Table 5). Comparative analysis was performed to evaluate the biochemical pregnancy rate in patients SARS-CoV-2 infection within 14 days after embryo transfer. The infected group showed a slightly lower biochemical pregnancy rate compared to the uninfected group (65.9% vs 58.1%). However, this difference was not statistically significant (p=0.081). Subsequently, logistic regression model 1 was adjusted for confounding factors, including fertilization type, type of transferred embryos, and number of high-quality embryos transferred and the adjusted odds ratio (OR) was 0.775 (95% CI, 0.526-1.141). To sum up, the results indicated that SARS-CoV-2 infection with 14 days may have no significant effect on the biochemical pregnancy rate.

**Table 4.** Pregnancy outcomes of univariate analysis.

| Outcomes | Non-COVID-19 | COVID-19 | P-value |
| --- | --- | --- | --- |
| <sup>a</sup> Biochemical pregnancy rate | 475 (65.9) | 79 (58.1) | 0.081 |
| <sup>b</sup> Implantation rate | 506/1150 (44.0) | 108/295 (36.6) | 0.181 |
| <sup>b</sup> Clinical pregnancy rate | 399 (58.2) | 84 (49.1) | 0.033 |
| <sup>c</sup> Early miscarriage rate | 10/107 (9.3) | 33/375 (8.8) | 0.861 |

Note:
<sup>a</sup>≤14days: biochemical pregnancy rate (infected group: 136, uninfected group: 721)
<sup>b</sup>≤28days: clinical pregnancy rate, implantation rate (infected group: 171, uninfected group: 686)
<sup>c</sup>≤10 weeks: early miscarriage rate (infected group: 696, uninfected group: 161):
Biochemical pregnancy rate, clinical pregnancy rate, and early miscarriage rate were presented as frequency (n) and percentages (%), while the embryo implantation rate is expressed as a rate.
$P < 0.05$ was considered statistically significant.

**Table 5.** Pregnancy outcomes of multivariable logistic regression.

| Outcomes | Unadjusted OR<br>(95%CI) | P-value | Adjusted OR<br>(95%CI) | P-value |
| --- | --- | --- | --- | --- |
| <sup>a</sup> Biochemical pregnancy rate | 0.718 (0.494-1.043) | 0.082 | 0.775 (0.526-1.141) | 0.196 |
| <sup>b</sup> Clinical pregnancy rate | 0.694 (0.496-0.972) | 0.033 | 0.736 (0.518-1.046) | 0.087 |
| <sup>c</sup> Early miscarriage rate | 0.936 (0.445-1.967) | 0.861 | 0.907 (0.414-1.986) | 0.808 |

Note:
<sup>a</sup> Logistic regression Model 1: fertilization type, embryo type, number of high-quality embryos transferred
<sup>b</sup> Logistic regression Model 2: fertilization type, embryo type, number of high-quality embryos transferred
<sup>c</sup> Logistic regression Model 3: FSH, AMH, COS protocol
95%CI, confidence interval; OR, odds ratio

In the second stage, clinical pregnancy rate and implantation rate were assessed. There was no statistically significant difference in the implantation rate between the infected group and uninfected group (36.6% vs. 44.0%, p=0.181). Interestingly, the clinical pregnancy rate showed statistically significant, the uninfected group and the infected group 58.2% and 49.1%, respectively, with a p-value of 0.033. However, after applying logistic regression model 2 to adjust for confounding factors (fertilization type, type of transferred embryos, and number of high-quality embryos transferred), this increased risk was no longer statistically significant (adjusted OR, 0.736, 95% CI, 0.518-1.046). In conclusion, SARS-CoV-2 infection within 28 days after embryo transfer may have not negatively affect the implantation rate and clinical pregnancy in patients.

In the third stage, the early miscarriage rate was 10/107 (9.3%) in the infected group and 33/375 (8.8%) in the uninfected group, with a p-value of 0.861. This suggested that the difference in early miscarriage rate between the two groups was not statistically significant. Logistic regression model 3 was further conducted to analyze the impact of SARS-CoV-2 infection within 10 weeks after embryo transfer. After controlling for the effects of three confounding factors (AMH, FSH, and COS protocol), the adjusted OR was 0.907 (95% CI, 0.414-1.986). This demonstrated that SARS-CoV-2 infection within 10 weeks after embryo transfer had not negative impact on the early miscarriage rate.

## Discussion

Infertility affects 8-12% of couples of reproductive ages, impairing their ability to conceive and negatively impacts their physical and mental well-being. At the same time, it is time-sensitive and gets worse with age. However, the COVID-19 pandemic has profoundly disrupted fertility services in most countries^8,23-25^. Despite advancements in our knowledge over the past three years, the impact of COVID-19 on male and female fertility, pregnancy, and fetal development in the field of ART remains largely unexplored^26^. ART specialists face challenges due to the lack of concrete scientific evidence needed for safe practice in the future. Meanwhile, unlike fertile couples who can naturally decide to conceive, infertile couples find themselves in a state of uncertainty, and unable to exercise their reproductive autonomy^27^.

Therefore, to incorporate new evidence to enhance the understanding of COVID-19 in relation to ART/IVF-generated pregnancies, benefiting both patients and clinicians, we analyzed 857 female patients underwent IVF/ICSI-ET treatment. It was the peak period of the epidemic in China from the end of 2022 to the beginning of 2023. To our best knowledge, this was the first report to investigate the impact of SARS-CoV-2 infection occurring at different time stages after embryo transfer on pregnancy outcomes in patients undergoing IVF/ICSI treatment.

Previous studies have indicated that the male and female reproductive systems are susceptible to SARS-CoV-2 infection. Negative effects on human semen parameters and male reproductive potential have been observed in some studies, but it seems that the research conclusions are controversial^5-8,28,29^. However, no significant impact of COVID-19 on ovarian reserves, ovarian function, follicular fluid parameters, or fertility has been reported^9-11^. Furthermore, when the male partner had a history of SARS-CoV-2 infection and later underwent IVF/ICSI-ET, they experienced similar rates of biochemical pregnancy rate, clinical pregnancy rate, implantation rate, and early miscarriage rate compared to non-infection couples^28^. However, in frozen embryo transfer (FET) cycles of patients with previous SARS-CoV-2 infection, in which oocytes were retrieved prior to infection, decreased pregnancy rates were observed, particularly in patients who recovered less than 60 days prior to embryo transfer^19^. On the other hand, females who underwent IVF/ICSI cycles and had a history of asymptomatic or mild SARS-CoV-2 infection showed similar outcomes in terms of embryo laboratory and IVF outcomes^9,17,30^. Nonetheless, inconclusive evidence suggests the need for further studies with large sample sizes.

For this study, we only included couples SARS-CoV-2 non-infection before oocyte retrieval and semen collection. We excluded female patients with a BMI below 18 or above 30 kg/m^2^, as well as those aged below 20 years or above 39 years. This is one of the advantages of our study, made possible by China’s government strict regulations in mitigating the adverse impacts of the COVID-19 pandemic on health before December of 2022. Consequently, we were able to focus on the effects of SARS-CoV-2 infection on pregnancy outcomes in couples undergoing IVF/ICIS-ET, excluding any confounding factors related to infected gametes. Interestingly, as mentioned earlier, we did not observe any significant impacts of SARS-CoV-2 infection on pregnancy outcomes, such as biochemical pregnancy, clinical pregnancy and implantation rate. A recent published multicenter prospective cohort study also reported that SARS-CoV-2 infection did not have an obvious effect on early human embryo development^14^. Considering the uncertainty regarding uterine receptivity alterations following infection, we suggest that SARS-CoV-2 infection occurring at different time stages after embryo transfer does not have a noticeable influence on pregnancy outcomes in patients undergoing IVF/ICSI treatment, as intact embryo development and uterine receptivity are maintained.

Several studies in general population showed pregnant women with COVID-19 are more likely to deliver preterm and have a substantially increased risk of preeclampsia, cesarean delivery, postnatal depression, neonatal complications, maternal mortality, fetal distress, perinatal death than uninfected women^31-41^. Furthermore, newborns of pregnant women exposed to SARS-CoV-2 may also face direct or indirect adverse health risks^42^. However, a Spain study reported that the overall incidence of pregnancy complications among women with SARS-CoV-2 infection was similar to uninfected women^43^. These findings suggest that the severity of COVID-19, the presence of COVID-19 variants, or COVID-19 vaccination may have varying impacts on pregnancy outcomes^44,45^. However, there is limited research exploring the effects of SARS-CoV-2 infection on women undergoing the ART treatment. One study shown that between the pre-COVID and COVID-19 periods in one of the earliest and most severe pandemic areas, the first trimester of ART pregnancy does not increase the risk of miscarriage^46^. In our study, we observed a slightly higher early miscarriage rate in infection patients than non-infected controls, but it was not reach statistically significant. However, owning to the limited follow-up time, we did not obtain longer-term pregnancy outcomes. While Calvo’s study observed high rates of surgical delivery among pregnant women admitted to the hospital with prenatal SARS-CoV-2 infection receiving ART^47^, and other evidence suggests that women infected with COVID-19 in ART-treated patients have a higher incidence of adverse obstetric and neonatal complications compared to women who conceive naturally^48^. Further studies are needed to confirm whether the combination of COVID-19 disease and IVF/ICSI treatment has a more negative impact on pregnancy fetal outcomes.

## Limitations

Our study has certain limitations. Firstly, due to a short follow-up period, the impact of SARS-CoV-2 infection on live birth and neonatal outcomes after embryo transfer remains uncertain. Secondly, Secondly, our analysis only included 857 participants who underwent fresh embryo transfer, and no cases of frozen-thawed embryo transfer were included. Therefore, a larger-scale, multicenter, long-term investigation is needed in the future to validate and monitor the long-term pregnancy outcomes and offspring health.

## Conclusions

The study’s findings indicate that SARS-CoV-2 infection within 10 weeks after embryo transfer may have a negative effect on the biochemical pregnancy rate, implantation rate, clinical pregnancy rate, and early miscarriage rate in patients. Moreover, it provides valuable insights, alleviating current concerns and shedding light on the actual risk of SARS-CoV-2 infection following embryo transfer. This research offers new perspectives on the real risk of COVID-19 infection after embryo transfer.

## Article Information

### Author Contributions

Xue-Fei Li, Yong-Jia Zhang drafted the original manuscript; Yong-Jia Zhang, Ming-Xing Chen and Ying-Ling Yao conducted data analysis. Yu-Bin Ding and Qi Wan conceived the study design and manuscript revision. Zhao-Hui Zhong, Xiao-Jun Tang, Xing-Yue Hu, Li-Juan Fu, and Meng-Di Wang were involved in the design of the questionnaire, data collection, and manuscript revision. Xin Luo, Li-Hong Geng, Li-Li Wang, and Xing-Yu Lv provided clinical consultation and assisted in data collection.

### Funding

This work was supported by the National Natural Science Foundation of China (82171664, 81971391) and Natural Science Foundation of Chongqing Municipality of China (CSTB2022NS CQ-LZX0062).

### Conflict of Interest Disclosures

We declare no competing interests.

## Data Availability

All data produced in the present study are available upon reasonable request to the authors.Request for access to the data should be made to the corresponding author at

## Acknowledgement

We are grateful to acknowledge all the participants of this study and the medical staff for their contribution to this work.

## Data sharing

Request for access to the data should be made to the corresponding author at. Data could be made available provided the applicant has appropriate ethics and author approval.

## Notes

### Competing Interest Statement

The authors have declared no competing interest.

### Author Declarations

the Ethics Committee of Sichuan Jinxin Xinan Women & Children's Hospital and the Ethics Committee of Chongqing Medical University

