## supplementary material for "The Impact of Post Embryo Transfer SARS-CoV-2 Infection on Pregnancy in In Vitro Fertilization: A Prospective Cohort Study"

### Questionnaire on COVID-19 and Assisted Reproduction

Number：□□□□□□

Medical record number：□□□□□□

patient name：________________

Hello! In order to investigate the impact of COVID-19 on the outcomes of assisted reproduction pregnancies, aiming to improve pregnancy rates and enhance pregnancy outcomes, we kindly ask you and your partner to carefully read and complete this questionnaire. Rest assured that the information provided will be kept strictly confidential. Thank you for your cooperation!

1. Have you been infected with COVID-19?

   ① Yes ② No

2. Date of COVID-19 infection：______Year___Month___Day

3. How were you diagnosed as COVID-19 positive?

   ① Nucleic acid testing ② Antigen testing

4. Did you experience symptoms during the course of your COVID-19 infection? (Select "Yes" to proceed to Question 5, select "No" to proceed to Question 6)

   ① Yes ② No

5. Which symptoms did you experience during your COVID-19 infection? (Select multiple options)

   Fever: Low-grade (37.3~38℃) □ Moderate (38.1~39℃) □ High-grade and above (39.1℃ and above) □

Fatigue ☐

Muscle aches ☐

Headache ☐

Joint pain ☐

Cough ☐

Nasal congestion☐

Chest pain ☐

Shortness of breath ☐

Loss of smell ☐

Loss of taste ☐

Diarrhea ☐

Decreased cognitive ability ☐

Sleep disturbances ☐

Depression or anxiety ☐

Decreased attention span ☐ Other____________(Specify)

6. Your COVID-19 vaccination status

6.1 Have you received the COVID-19 vaccine? (Select one option)

① Yes ② No (If "Yes," proceed to Question 6.2; if "No," proceed to Question 7)

6.2 If vaccinated, please indicate the type of vaccine received (Select one option)

① Inactivated vaccine (a, Beijing Biological Products; b, Wuhan Biological Products; c, Sinovac Biotech-Beijing Kexing; d, Chengdu Biological Products; e, Changchun Biological Products; f, Shanghai Biological Products; g, Lanzhou Biological Products) (Select ① to proceed to Questions 6.3.1 and 6.3.2)

② Recombinant subunit vaccine (Select ② to proceed to Question 6.3)

③ Adenovirus vector vaccine (Select ③ to proceed to Question 6.3.1)

(Inactivated vaccines generally require 2 doses, recombinant subunit vaccines require 3 doses, and adenovirus vector vaccines require 1 dose)

6.3 If vaccinated, please provide the vaccination dates: (Select the dates using the sliding scale)

6.3.1 Date of the first dose: Year______ Month______ Day______

6.3.2 Status of the second dose: a, Received; Date of vaccination: Year______ Month______ Day______ b, Not received (Select one option)

6.3.3 Status of the third dose: a Received, Date of vaccination: Year______ Month______ Day______ b Not received (Select one option)

6.4 Did you experience any adverse reactions within one week of receiving the vaccine? (Select one option)

① None ② Yes (If "Yes," proceed to Question 6.4.1)

6.4.1 If yes, please select the adverse reactions experienced (Select multiple options)

Fever ☐

Chills ☐

Fatigue ☐

Muscle aches ☐

Joint pain ☐

Headache dizziness ☐

Nausea or vomiting ☐

Diarrhea ☐

Redness at the injection site ☐

Swelling at the injection site ☐ Other__________ (Specify)

7. Did you use any medication for treatment during the infection process? (If "No," skip Question 8)

① Yes ② No

8. If you used medication, please specify the type(s) used. (Select multiple options)

Acetaminophen □

Ibuprofen □

Lianhua Qingwen granules/capsules □

Jinhua Qinggan granules □

Antiviral granules □

Huoxiang Zhengqi liquid □

Banlangen granules □ Other__________ (Specify)

| Note: |
| --- |

Investigator：____________ Date of survey: _______ Year_____Month____Day

Reviewer：______
